# Vasoplegic syndrome in cardiovascular surgery; evaluating effects of Sevoflurane and Glibenclamide in a porcine model

**DOI:** 10.1101/2023.04.17.23288708

**Authors:** Andreas Winter, Pascal Nepper, Marcus Hermann, Franziska Bayer, Stephanie Rieß, Spiros Marinos, Arnaud Van Linden, Tomas Holubec, Andrea Steinbicker, Kai Zacharowski, Thomas Walther, Fabian Emrich

## Abstract

**Background:** Vasoplegic syndrome is frequently observed during cardiac surgery and resembles a complication of high mortality and morbidity. There is a clinical need for therapy and prevention of vasoplegic syndrome during complex cardiac surgical procedures. Therefore, we investigated different strategies in a porcine model of vasoplegia.

**Methods:** We evaluated new medical therapies and prophylaxis to avoid vasoplegic syndrome in a porcine model. After induction of anesthesia, cardiopulmonary bypass was established through median sternotomy and central cannulation. Prolonged aortic cross-clamping (120 min) simulated a complex surgical procedure. The influence of sevoflurane-guided anesthesia (sevoflurane group) and the administration of glibenclamide (glibenclamide group) were compared to a control group, which received standard anesthesia using propofol. Online hemodynamic assessment was performed using PiCCO^®^ measurements. In addition, blood and tissue samples were taken to evaluate hemodynamic effects and the degree of inflammatory response.

**Results:** Glibenclamide was able to break through early vasoplegic syndrome by raising the blood pressure and systemic vascular resistance as well as less need of norepinephrine doses. Sevoflurane reduced the occurrence of the vasoplegic syndrome in the mean of stable blood pressure and less need of norepinephrine doses.

**Conclusion:** Glibenclamide could serve as a potent drug to reduce effects of vasoplegic syndrome. Sevoflurane anesthesia during cardiopulmonary bypass shows less occurrence of vasoplegic syndrome and therefore could be used to prevent it in high-risk patients.

**Clinical Perspective; what is new?:**

- to our knowledge, this is the first randomized in vivo study evaluating the hemodynamic effects of glibenclamide after the onset of vasoplegic syndrome
- furthermore according to literature research, there is no study showing the effect of sevoflurane-guided anesthesia on the occurrence of a vasoplegic syndrome

**Clinical Perspective; clinical implications?:**

- to achieve better outcomes after complex cardiac surgery there is a need for optimized drug therapy and prevention of the vasoplegic syndrome

---

Despite the frequent use of minimally invasive approaches, conventional cardiopulmonary bypass [CPB] and cardiac arrest remain the routine techniques in complex surgical procedures.

CPB may be associated with the occurrence of the vasoplegic syndrome [VS], which leads to higher morbidity and mortality. VS is characterized by a vasodilatation and reduced systemic vascular resistance [SVR], leading to a contributive shock with the need for catecholamines and impaired peripheral perfusion ^1^.

VS is reported to occur in up to 20 to 63% of patients, ^2, 3^ and may be related to longer CBP duration ^4^. Current therapy includes the administration of volume and vasopressors, which in turn are associated with increased mortality and morbidity ^1, 3, 5^. Further possibilities of drug therapy are methylene blue and vasopressin ^6, 7^.

The pathogenesis is considered to be multifactorial. Contact activation, adenine triphosphate [ATP] deficiency, activation of the complement and coagulation systems as well as various anesthetics and their effects on vascular tone are discussed ^1, 3, 5, 8^.

This study aims to evaluate new medical approaches to avoid and reduce VS in a porcine model with prolonged CPB. The influence of two different concepts of anesthesia were compared: total intravenous anesthesia [TIVA] using propofol and volatile anesthetics [VA] by means of sevoflurane. Due to a lack of application devices VA is not commonly used during CPB while TIVA is the most used anesthetic procedure. Positive effects of sevoflurane regarding the hemodynamic effects were previously seen in different studies ^8, 9^.

The pathophysiology of the VS is very similar to that of a systemic inflammatory response syndrome [SIRS]. A cytokine storm, which happens in SIRS by contact with exogenous pathogens, also occurs in VS after CPB. Here, however, contact activation is the origin of the immune reaction. In both cases, the release of cytokines in conjunction with the expression of nitric oxide [NO] leads to peripheral vasodilation and loss of endothelial integrity ^10, 11^. A measurement of pro-inflammatory cytokines, interleukin-1 beta [IL-1β], interleukin 6 [IL6], and the tumor necrosis factor alpha [TNFα] can therefore quantify an immune reaction (Groeben et al. 2005). The anti-inflammatory interleukin-10 [IL-10] might play a role in the cascade following SIRS ^12^.

Hemodynamic data, specifically measurement of the mean arterial pressure, SVR and the catecholamine requirement serve as the primary indicator of occurrence of a VS. Another marker is free NO and the expression of endothelial NO synthase [eNOS] as well as the inducible NO synthase [iNOS] in tissue samples from smooth vascular muscles. By increasing the concentration of cyclic guanine monophosphate [cGMP], NO leads to relaxation of the smooth vascular muscles and vascular dilation^13, 14^.

Another mechanism associated with the development of pathological vasodilation is the activation of ATP sensitive potassium channels [K_ATP_]. A reduced concentration of ATP in the case of ischemic malperfusion means that potassium channels are no longer inhibited and are therefore passively activated. This also leads to relaxation of the smooth vascular muscles.

A sulfonylurea called glibenclamide, which is already approved as an antidiabetic agent and currently investigated in a phase II study as an intravenous agent against cerebral edema ^15, 16^, inhibits this K_ATP_ and could therefore attenuate the VS ^17, 18^.

A known effect of the volatile anesthetic sevoflurane is the inhibition of the activation of neutrophil granulocytes. This leads to a reduced release of PNM elastase and MAC-1 expression and therefore could prevent a VS ^11, 19^.

## Methods

The data that support the findings of this study are available from the corresponding author upon reasonable request.

### Porcine model of vasoplegic syndrome

We established an animal model for VS in adherence to the European directive 2010/63/EU with a positive vote from the regional council Darmstadt of June 6^th^ 2022 under the file number FK/2036. In this porcine model, 38 German female landrace pigs, 6 months old, with a body weight of 74.6±1.5 kg were examined. An initial pilot study to establish the experimental setup was performed on five animals. These were excluded from further analysis as well as three animals due to procedural complications (myocardial ischemia, n=1) and a toxic effect of incorrectly stored study drug (n=2). The remaining 30 pigs were divided in 3 Groups with randomized adjudication of testing:

- Routine propofol anesthesia as a control group [CG]

- Sevoflurane [SG]

- Propofol + glibenclamide [GG].

The study drug glibenclamide (Glybenclamid G0639, Sigma-Aldrich) was administered 45 min after weaning from cardiopulmonary bypass [post-CPB] as bolus of 10 mg/kg at a rate of 500 mg/min. This bolus was directly followed by a continuous infusion via syringe pump in a dosage of 10 mg/kg/h. Glibenclamide was dissolved with 100% dimethyl sulfoxide (DMSO, D8418, Sigma-Aldrich) at a concentration of 100 mg/ml. Hemodynamic effects of dimethyl sulfoxide without glibenclamide were ruled out by administering it in four animals in CG and SG before sacrifice. A previous study had a similar observation ^20^. The experiments were carried out in the central animal research facility of the Johann Wolfgang Goethe University Hospital in Frankfurt am Main, Germany.

### Induction and maintenance of anesthesia

Premedication was carried out via intramuscular application of 20 mg/kg ketamine (Ketaset^®^, Zoetis), 1 mg/kg xylazine (Rompun^®^, Elanco) and 0.5 mg/kg midazolam (Midazolam-hameln, Hameln-Pharma). During pre-oxygenation with a nose cone, intravenous cannulation (20G Braunüle^®^, Braun) of the lateral auricular vein was performed. Anesthesia was then induced by intravenous bolus administration of 1 mg/kg propofol (Propofol, MCT Fresenius^®^) in the CG and GG, 3-5 µg/kg Fentanyl (Fentanyl^®^, Piramal) and 0.1 mg/kg pancuronium (Pancuroniumbromid, Panpharma). After orotracheal intubation in prone position, volume-controlled ventilation was initiated (Sulla 808-V, Dräger), aiming at a tidal volume of 8 ml/kg body weight and a positive end-expiratory pressure of 5 cmH_2_O. A physiological end-expiratory concentration of carbon dioxide [CO_2_] was monitored by capnography (Vamos^®^, Dräger) connected to the oral tube. Anesthesia was maintained through continuous application of 0.3 µg/kg/min fentanyl and 10 mg/kg/h propofol via two syringe pumps (Perfusor^®^ Space, Braun) in the CG and GG. In the SG, anesthesia was maintained and induced via gas insufflation using a Vapor (Vapor^®^, Dräger) and fentanyl administration. The depth of anesthesia was measured via the end-expiratory minimum alveolar concentration [MAC], aiming at 0.8-1.0 (Volume concentrations of 2.4-2.6 %) using the Vamos^®^ device. Repetitive doses of 0.02 mg/kg pancuronium caused an overlapping relaxation. For basal volume substitution, a balanced full electrolyte solution ran continuously at a rate of 100 ml/h via an infusion pump (Infusomat^®^ Space, Braun).

### Monitoring

Continuous basic monitoring included an electrocardiogram [ECG], measuring heart rate [HR], peripheral oxygen saturation and a rectal temperature probe. Following supine positioning of the animal, a central venous catheter (Arrow, 7 F) was introduced percutaneously into the external jugular vein for drug application, thermodilution [TD] measurements and central venous pressure [CVP] measurements. Invasive blood pressure and extended hemodynamic monitoring was established via the femoral artery using the PiCCO^®^ system (PiCCO ^®^ Pulsion Medical Systems). Both catheters were inserted under sterile conditions.

A Foley catheter was introduced transurethral (CARE Flow 16 Ch., GHC).

### Surgical procedure

Prior to the supra-sternal skin incision, anesthesia was deepened with a bolus application of 3-5 µg/kg fentanyl and 0.02 mg/kg pancuronium. Surgery was performed under sterile conditions

After sternotomy, heparin (Heparin-Natrium, Braun) was administrated at a dose of 500 I.E./kg intravenously, aiming at an activated clotting time of >450 sec. The ascending aorta (Arterial Cannula 20 Fr, Andocor) and the right atrium (Venous Cannula, 32 F–24 F, L 380 mm–42 mm, Sorin Group) were cannulated according to local standards. An aortic root vent was then placed for intergrade delivery of cardioplegia and venting of the heart. CPB was initiated using the heart-lung machine [HLM] model MAQUET HL 20 (Maquet). Extracorporeal blood flow was adjusted to maintain a cardiac index [CI] of at least 2.4 ml/min/kg body weight. Then aortic cross-clamping [ACC] was performed and 1000ml of Del Nido cardioplegic solution (4:1 crystalloid:blood; Pharmacy University Frankfurt) was infused in a flow and pressure controlled manner. Cardiac arrest was achieved under hypothermic conditions. MAP during CPB was maintained between 50-70 mmHg. During ACC, lung ventilation was paused. In the SG, sevoflurane administration was performed through the oxygenator (CAPIOX® FX25RE, Terumo). Vamos^®^ measured concentration in the outflow tube. After 60 min of ACC another 500ml of Del Nido cardioplegic solution was infused through the aortic root cannula. Blood gas and hemoglobin levels were measured online. Gas flow was adjusted to post oxygenator CO_2_ measurements. A passively mild hypothermia occurred until 35°C. After 60 min, reheating was started to regain normothermy until the end of ACC.

After 120 min of ACC, mixed venous reperfusion of warm blood (“hot shot”) for 3 min was carried out through the aortic root cannula, and then ACC was released. In case of ventricular fibrillation, defibrillation with internal paddles was performed (n=10). Ventilation of the lungs was commenced after recruitment maneuver. After 60 min of reperfusion, animals were weaned off CPB.

In the next 90 min of post-CPB observation, hemodynamic monitoring was performed via MAP and necessary norepinephrine doses. In the GG, the study drug was infused intravenously 45 min after weaning from CPB. During the post-CPB observation, anesthesia in SG was again maintained via tracheal gas insufflation of sevoflurane.

The experiment was finalized after 90 min of post-CPB. 15ml of T61 (tetracainhydrochlorid, membezoniumiodid, embutramid; Intervet) was given intravenously to sacrifice the animal.

### Hemodynamic data

The study was divided into four time periods:

**Surgical procedure:** time until the start of cardiopulmonary bypass and aortic cross-clamping

**ACC:** time on cardiopulmonary bypass while the aorta is cross-clamped (120min)

**CPB:** time on cardiopulmonary bypass during reperfusion (60 min)

**Post-CPB:** time after cardiopulmonary bypass for hemodynamic observations (90 min)

Through the PiCCO^®^ system, hemodynamic parameters were measured either through thermodilution [TD] or through pulse contour analysis [PCA]:

Cardiac index [CI_TD_] and [CI_PCA_]

Extravascular lung water index [ELWI]

Global end-diastolic volume index [GEDI]

Global ejection fraction [GEF]

Left ventricular contractility [dPmx]

Systemic vascular resistance index [SVRI]

With the initiation of CPB, the hemodynamic parameters were documented every 15 min, including MAP, arterial flow velocity on CPB and the current norepinephrine dose. In addition, the venous oxygen saturation, the pCO_2_ and pO_2_ as well as the current hemoglobin values were recorded every 30 min. HR was recorded every 15 min after ACC.

After weaning from CPB a closer monitoring of hemodynamic parameters was carried out and HR, MAP, CVP, dPmx and norepinephrine dose were then documented every 5 min until the end of the experiment. The cardiac index was also determined via PCA and recorded every 5 min. After 45 min post-CPB, the documentation interval was reduced again to one minute for a period of 10 min in CG and GG, to evaluate the effect of glibenclamide more closely. During post-CPB, PiCCO^®^ measurements via TD were carried out every 30 min to validate the continuous measurements.

Volume levels were held stable by administration of crystalloid solutions according to PiCCO^®^ measurements of GEDI and ELWI as well as CVP and HR before, during and after CPB.

### Laboratory testing

Blood samples were taken at baseline directly after establishment of the central venous catheter and at the end of the experiment shortly before sacrifice. Blood collection tubes were sent to the central laboratory at the University Hospital in Frankfurt. A secondary batch was centrifuged, pipetted off and frozen at -80°C for later analysis. In addition, a point of care blood gas analysis [BGA] (i-STAT^®^ Alinity, Abbot) and a blood sugar determination (ACCU-CHEK^®^, Roche) were carried out at four time points throughout the experiment.

In addition, serum creatinine, lactate dehydrogenase, creatine kinase (CK and CK-MB), total protein and albumin were measured. The thromboplastin time and the activated partial thromboplastin time were determined for coagulation diagnostics.

### Tissue samples

Samples were taken from the lungs, heart, liver, kidney, femoral artery, aorta and pulmonary artery directly after sacrificing the animals, snap-frozen in liquid nitrogen and stored at -80°C for further analysis.

### Sample processing

#### Blood samples

The concentration of the following parameters were determined using commercially available ELISA kits. IL-1β, IL-6, IL-10 and TNF-α were measured by Quantikine^©^ porcine ELISA Kits (R&D Systems). Concentrations of CD11b, PMN-elastase, malondialdehyde, eNOS and iNOS were measured by Kits from BIOZOL Diagnostica.

#### Aortic tissue samples

The samples were cut into 100 mg pieces, crushed and homogenized with 0.9 ml phosphate-buffered saline, centrifuged at 6000 rpm for 3×15 sec in appropriate vessels with dry ice cooling. Then the supernatant was pipetted off and again centrifuged at 5000 g for 5 min at 4°C. The ELISA was carried out from this supernatant. iNOS and eNOS concentrations from aortic tissue were measured using Quantikine^®^ ELISA Immunoassay kits.

#### Quantikine^®^ Elisa Immunoassay

The process of an ELISA is briefly explained using the R&D operating instructions ^21^. In order to achieve the most accurate results as possible, each measurement was carried out twice and the values were averaged.

### Statistical analysis

All results were expressed as the mean±standard error of the mean [SEM]. The statistical software SPSS v29 (IBM SPSS Statistics, IBM) was used to analyze all data.

All data was tested for normal distribution using the Shapiro-Wilk test per measurement point and group. If the distribution was normal, the test for homogeneity of variance was then carried out using the Levene test. Within 10 min after glibenclamide application, the Mann-Whitney-U test was used to test for significance between CG and GG. Significant differences within one group were analyzed either using a paired-samples t-test for normally distributed data or a Wilcoxon matched-pair test. For all other time stamps, all three groups were compared. With normal distribution and homogeneity of variance, an ANOVA with Bonferroni’s *post hoc* was used. Otherwise, difference between all groups were shown using the Kruskal-Wallis test with Dunn-Bonferroni correction.

Body surface area [BSA] was calculated according to the following formula: BSA= 0.0798×kg/body weight^2/3^ ^22^.

*P* values <0.05 were considered significant.

### Hematocrit [HCT] correction

Because of high priming volumes using CPB and high volume doses infused during the study for hemodynamic stability, there was a relevant dilution of the blood samples taken at the end of experiment. With the help of an HCT correction according to Schmid et al., all blood parameters which are given as concentrations, were multiplied by an HCT factor ^19^. This factor was obtained by dividing the HCT before the start of the surgical procedure by the HCT at the end of the experiment. Since the HCT could not be determined at all times when lactate and blood sugar were taken, these parameters are expressed without any correction.

## Results

All animals in the CG showed signs of the VS in the mean of higher norepinephrine doses maintaining the MAP and SVRI. The experiment therefore resembles a valid opportunity to examine the VS. In The SG Group no VS was observed. In GG VS could be reduced directly after application of glibenclamide.

### Control Group (CG)

In the CG VS occurred in post-CPB observation. MAP remained constantly low under norepinephrine therapy. SVRI dropped significantly during post-CPB observation, from 1459±135.7 dyn*sec*cm^-5^*m^2^ to 1187.5±93.7 dyn*sec*cm^-5^*m^2^ at the end of experiment (fig. 5;p<0.05). In order to keep MAP between 60 – 70 mmHg, the norepinephrine doses had to be adjusted from 0.12±0.02 µg/kg/min starting post-CPB observation to 0.373±0.035 µg/kg/min at the end of experiment (fig. 4;p<0.01). The heart rate raised from 102.8±6.1 bpm min starting post-CPB observation to 119.6±5.8 bpm at 45 min (fig. 2;p<0.01) and stayed at this level until the end of experiment. CI_PCA_ did not change significantly. DPmx raised significantly continuous over the time from 532±67.4 mmHg/sec at the beginning to 1043.7±123.8 mmHg/sec (p<0.01). There were no significant changes in CVP, GEDI, ELWI, and GEF during post-CPB observation in the CG.

**Figure 1:**
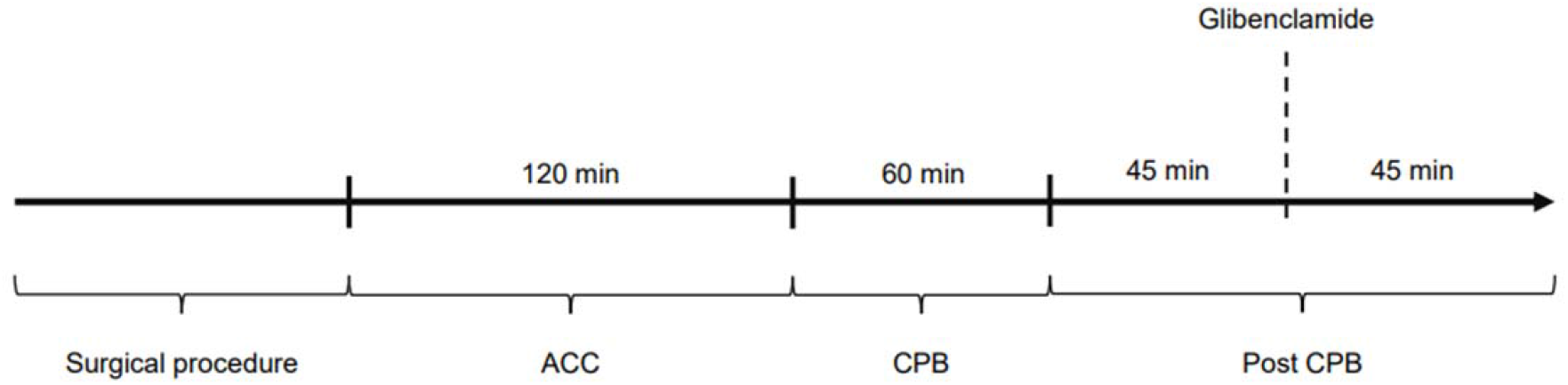
Experimental protocol of the study

**Figure 2.**
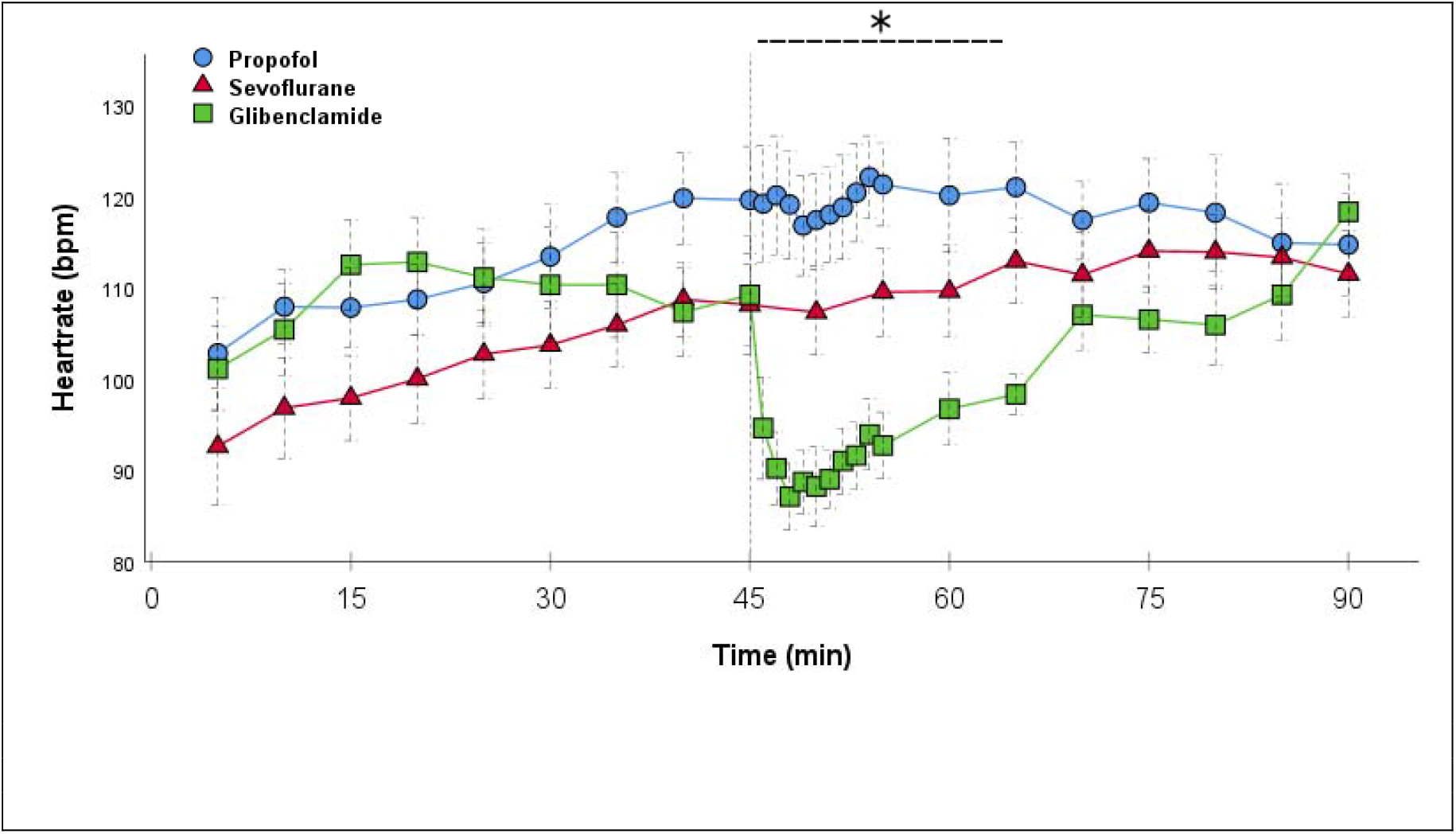
Heartrate: Continuous monitoring of the heartrate during post-CPB observation. Values are expressed as mean±SEM. * (p<0.05) represents significant differences between glibenclamide and control group at the respective time stamps (*ANOVA with Bonferroni’s post hoc*). The reference line at 45 min signals the start of glibenclamide application.

**Figure 3.**
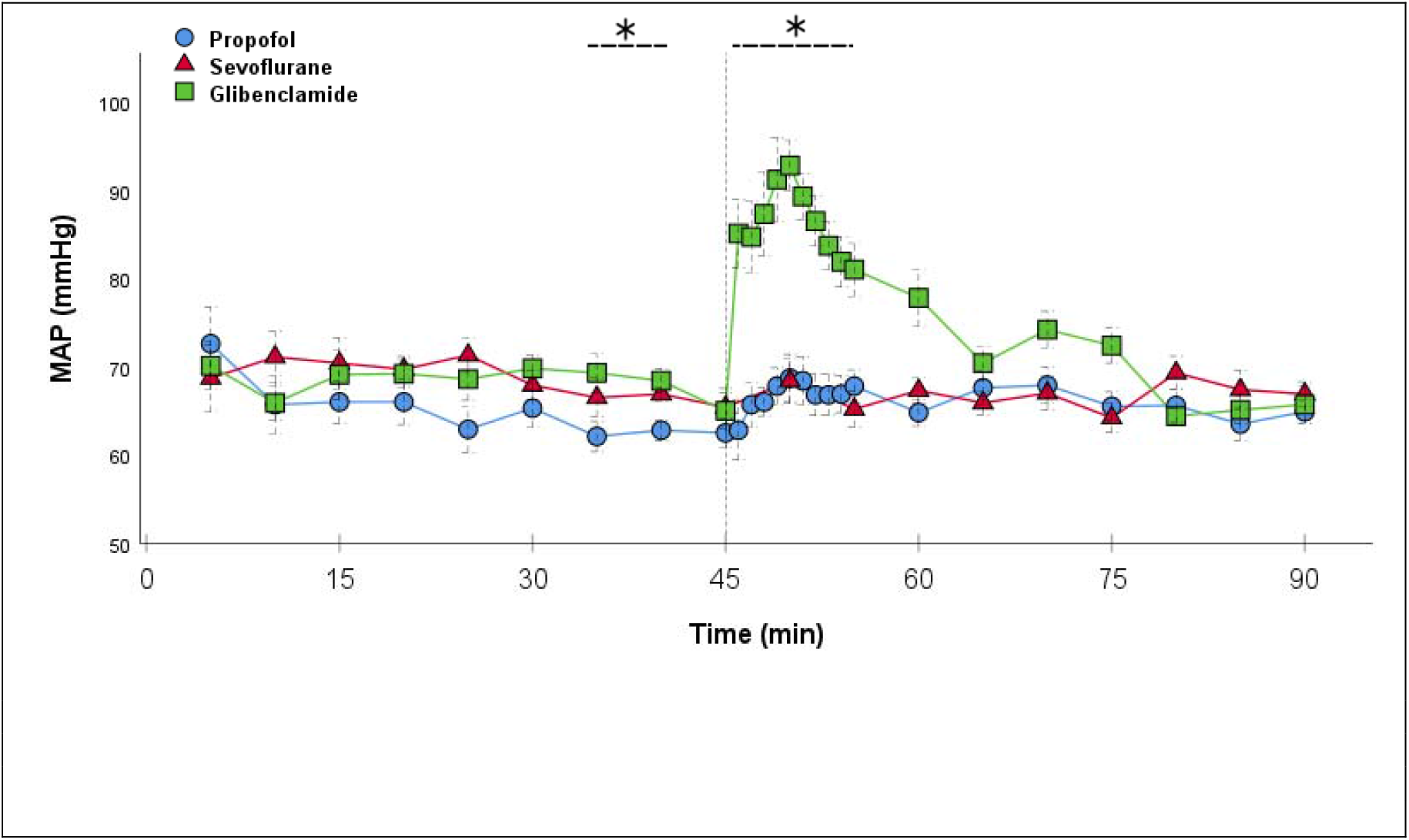
MAP: Continuous monitoring of the mean arterial pressure during post-CPB observation. Values are expressed as mean±SEM. * (p<0.05) represents significant differences between glibenclamide and control group at the respective time steps (*Kruskal-Wallis test with Dunn-Bonferroni correction*). The reference line at 45 min signals the start of glibenclamide application.

**Figure 4.**
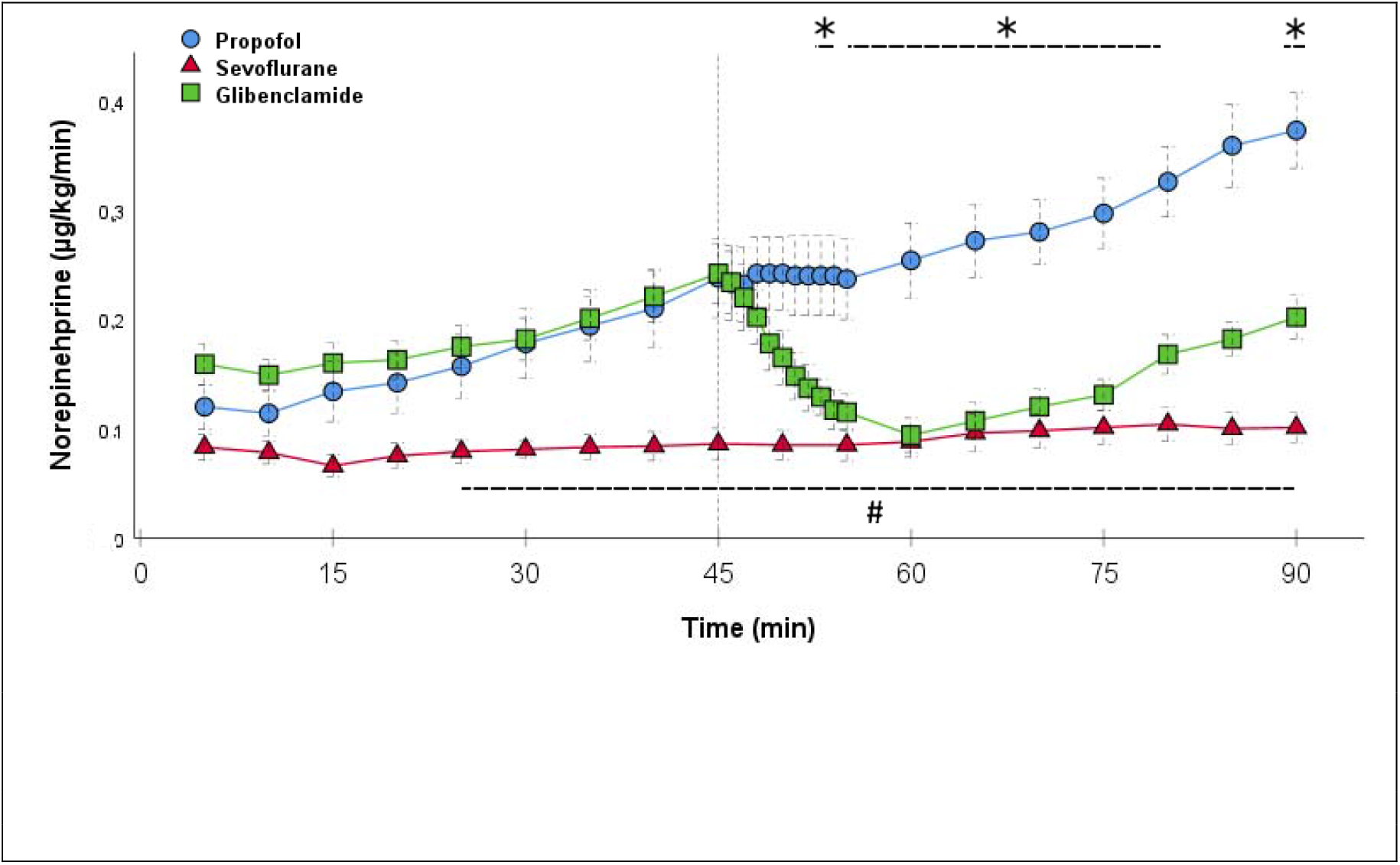
Norepinephrine: Continuous application of norepinephrine during post-CPB observation. Values are expressed as mean±SEM. * (p<0.05) represents significant differences between glibenclamide and control group at the respective time steps. # (p<0.05) represents significant differences between sevoflurane and control group at the respective time steps (*Kruskal-Wallis test with Dunn-Bonferroni correction)*. The reference line at 45 min signals the start of glibenclamide application.

**Figure 5.**
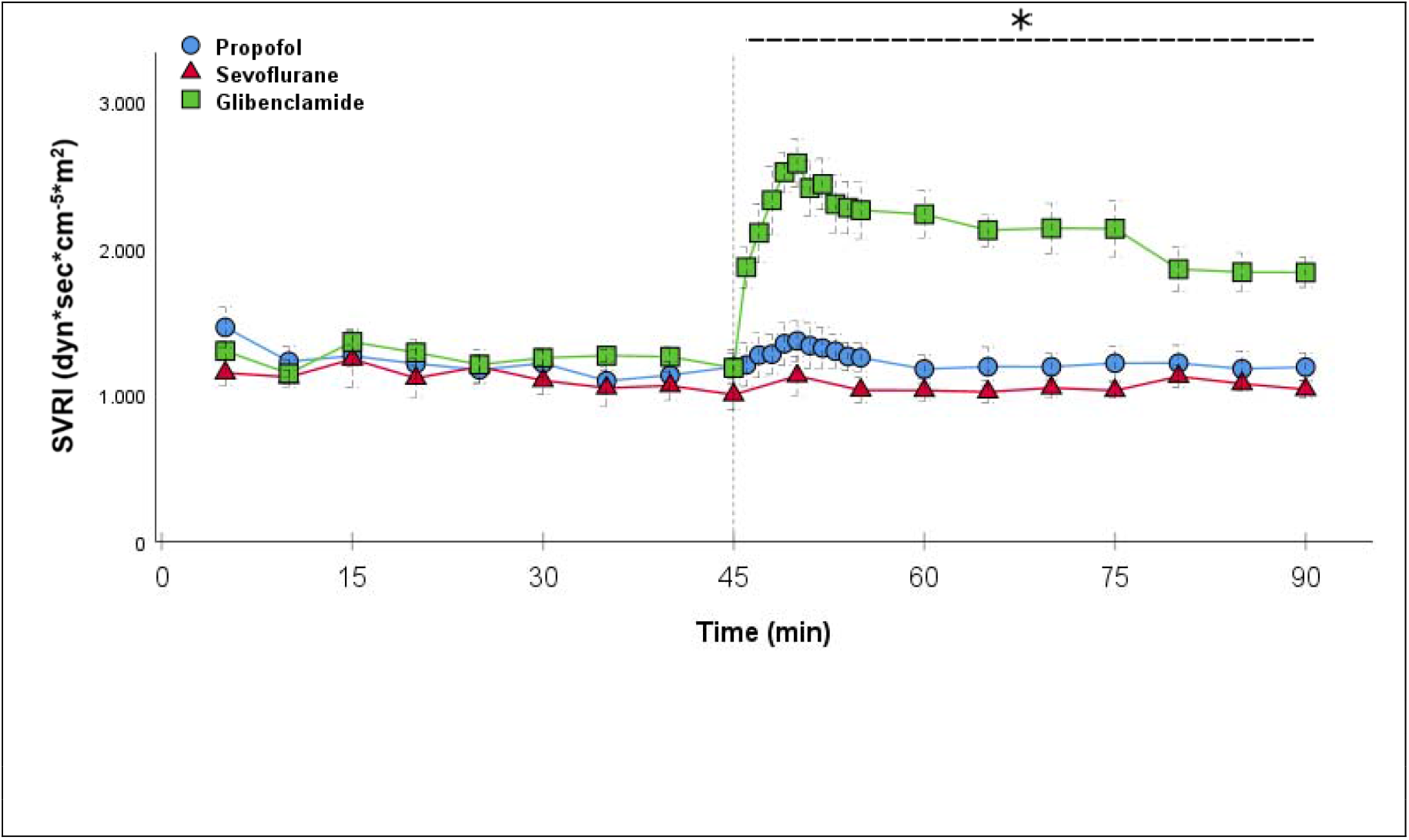
SVRI: Continuous monitoring of the systemic vascular resistance index during post-CPB observation. Values are expressed as mean±SEM. *(p<0.05) represents significant differences between glibenclamide and control group at the respective time steps (*Kruskal-Wallis test with Dunn-Bonferroni correction)*. The reference line at 45 min signals the start of glibenclamide application.

Serum-lactate stayed constant throughout the experiment, starting with 1±0.1 mmol/l and ending with 1.6±0.2 mmol/l (fig. 7;p=0.575). The concentration of all Cytokines raised compared pre and post-CPB. IL-1β raised from 30.4±4.5 pre-CPB to 64.2±9.1 pg/ml at the end (p<0.01), IL-6 raised from non-detecting concentrations (<2.03pg/ml) to 107.2±33.8 pg/ml, IL-10 from 30.2±1.9 pg/ml to 38.9 pg/ml (p<0.01), TNF-α from 63.8±6.2 pg/ml to 118±13.2 pg/ml (p<0.001). Malondialdehyde raised from 0.37±0.02 nmol/ml to 0.5±0.04 nmol/ml (p<0.05). PMN-Elastase raised from 2.5±0.1 ng/ml to 5.4±0.3 ng/ml (p<0.01)

### Glibenclamide (GG) vs. Propofol (CG)

Significant hemodynamic differences between the groups were seen in MAP, SVR, HR, CI and Lactate. After the application of glibenclamide at 45 min post-CPB, the HR dropped significantly from 109.2±6.5 bpm to 87.1±3.6 bpm within 3 min in GG compared to CG 119.1±5.9 bpm (fig. 2;p<0.001). Simultaneously, MAP increased from 65±2.1 mmHg to 92.8±2.9 mmHg in 5 min at 50 min post-CPB compared to CG 68.7±2.7 mmHg (fig. 3;p<0.01). Simultaneously, the necessary norepinephrine dose could be reduced from 0.242±0.027 µg/kg/min at 45 min post-CPB to 0.094±0.016 µg/kg/min 15 min after application at 60 min post-CPB compared to CG 0.254±0.035 µg/kg/min at 60 min post-CPB (fig. 4;p<0.01). The SVRI increased from 1182.8±103.8 dyn×sec×cm^-5×^m^2^ to 2576.9±164.2 dyn×sec×cm^-5×^m^2^ in 5 min at 50 min post-CPB compared to CG 1366.6±143 dyn*sec*cm^-5^*m^2^ at 50 min post-CPB (fig. 5;p<0.01). SVRI stayed significantly higher compared to CG until the end of the experiment. CI_PCA_ normalized from 4.1±0.3 l/min/m^2^ to 2.7±0.13 l/min/m^2^ within 4 min at 49 min post-CPB compared to CG 3.9±0.41 l/min/m^2^ at 49min post-CPB (fig. 6;p<0.05). Furthermore, the CI_PCA_ had its lowest value of 2.4±0.1 l/min/m^2^ at 65 min post-CPB compared to CG 4.4±0.5 l/min/m^2^ (p<0.01) and stayed significantly lower until the end of experiment. According to the CI, global Ejection fraction was reduced after application of glibenclamide 19.2±1.2 % vs. 26.9±1.8% at 60 min post-CPB, (p<0.05) and 18.4±0.89 % vs. 26.5±1.6 % at 90 min post-CPB. (p<0.05). No further significant hemodynamic differences were found through CVP, ELWI, GEDI and dPmx. Volume administration showed no significant differences between the groups. There was no effect of dimethyl sulfoxide in carrier observations.

**Figure 6.**
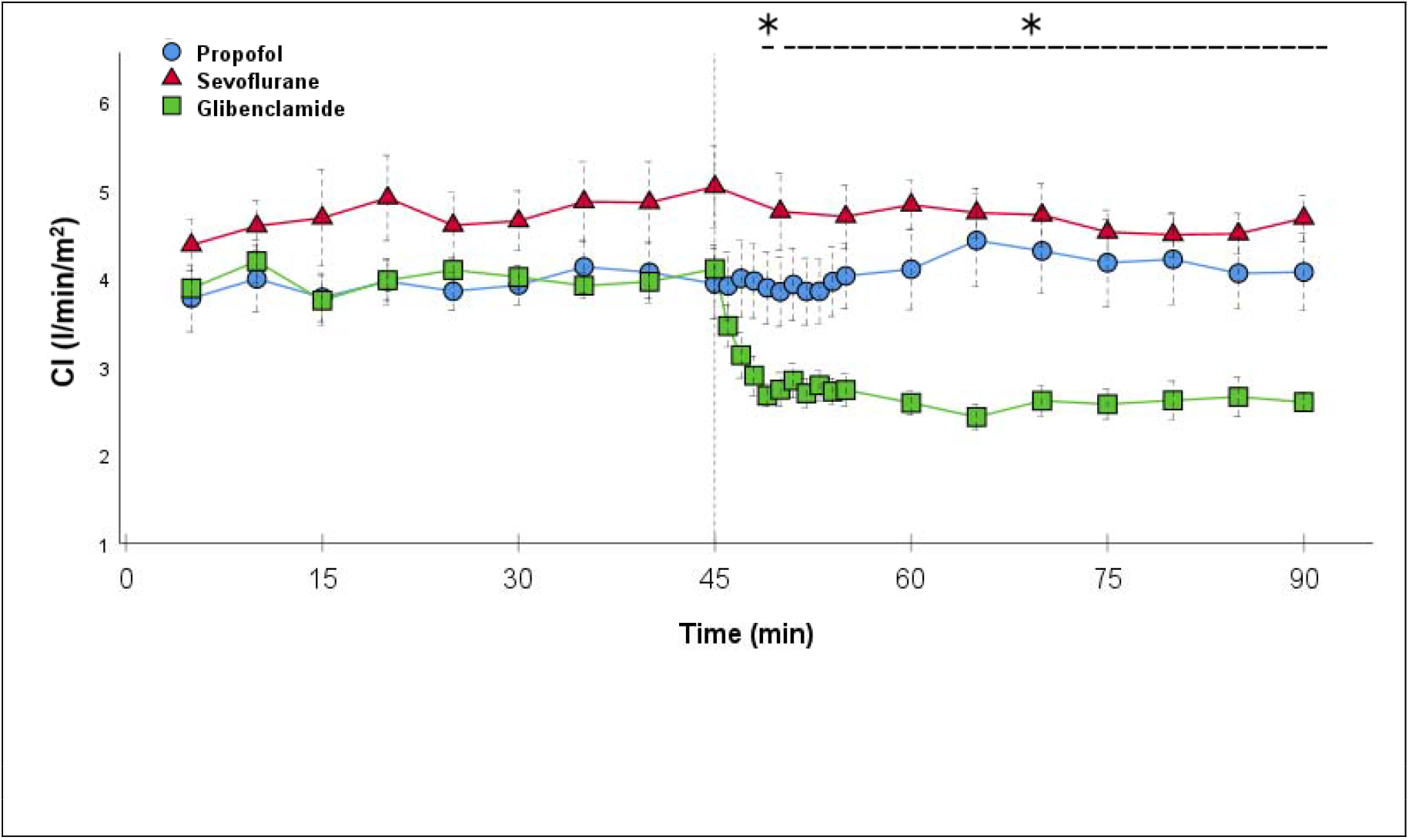
CI_PCA_: Continuous monitoring of the pulse contour cardiac index during post-CPB observation. Values are expressed as mean±SEM. * (p<0.05) represents significant differences between glibenclamide and control group at the respective time steps (*Kruskal-Wallis test with Dunn-Bonferroni correction)*. The reference line at 45 min signals the start of glibenclamide application.

**Figure 7.**
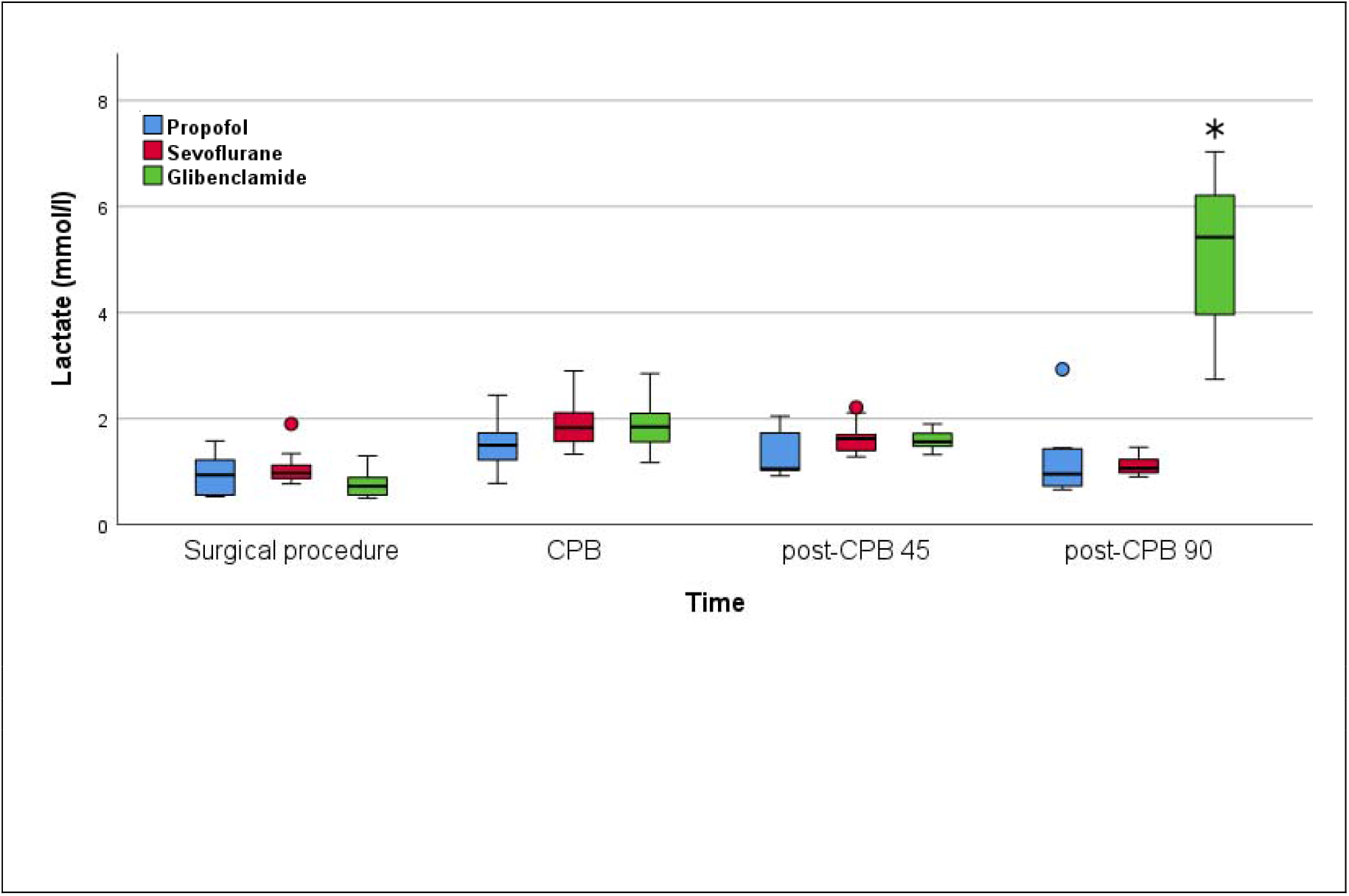
Lactate: Concentration of serum-lactate during experiment. Surgical procedure resembles baseline values. CPB resembles the time point 60 min during CPB time. Post-CPB 45 resembles the time point 45 min during post-CPB observation, right before glibenclamide application. Post-CPB 90 resembles the end of experiment. Values are expressed as boxplots, continuing the median as a horizontal line inside of the box. The first and third quantile are the edges of the box. Whiskers have a distance about 1.5 interquartile range to the edges. Outliers are plotted as dots. *(p<0.05) represents significant differences between glibenclamide and control group at the respective time steps (*Kruskal-Wallis test with Dunn-Bonferroni correction*).

Significant effects of laboratory measurements were seen in the Lactate concentration after application of glibenclamide, serum-lactate raised from 1.6±0.6 mmol/l to 5.2±0.4 mmol/l at the end of the experiment, 90 min post-CPB compared to CG 1.219±02 mmol/l (fig. 7;p<0.001). No further significant differences in central laboratory testing, ELISA or BGA were found especially in the concentration of LDH, Creatinine, CK, CK-MB, IL1β, IL6, IL-10, malondialdehyde, PMN-Elastase and CD11b.

### Sevoflurane (SG) vs. Propofol (CG)

Significant hemodynamic differences between the groups were observed regarding the dosages of norepinephrine. It showed lower levels beginning 25 min post-CPB 0.079±0.01µg/kg/min compared to CG 0.157±0.029µg/kg/min (fig. 4;p<0.05) until the end of the experiment 0.1±0.014µg/kg/min compared to CG 0.373±0.035 µg/kg/min (fig. 4;p<0.001). CI_PCA_ was not significantly higher in the SG at 45 min post-CPB 5.±0.5 l/min/m^2^ compared to CG 3.9±0.4 l/min/m^2^ (fig. 6;p=0,65), but at one time GEF was significant higher in the SG 29,6±0,8 % at 30 min post-CPB compared to CG 24. 8± 1.4 % (p<0.05).

Urine output was significant higher in the SG 3.9±0.4 ml/kg/h compared to CG 1.7±0.3 ml/kg/h at the end of the experiment (p<0,001).

Volume administration controlled via PiCCO^®^ data GEDI and ELWI showed no significant differences in between the groups. In comparison to the CG there were no significant differences in MAP, SVRI or other hemodynamic parameters.

There were significantly lower TNF-α levels at the end of experiment 78.8±7.5 pg/ml compared to CG 118±13.2 pg/ml (fig. 8;p<0.05). No further significant effects in central laboratory testing, ELISA or BGA were found. PMN-Elastase, Creatinine levels and other laboratory testing as described above had no significant differences in between all groups at the end of experiment (p=0.566;PMN, p=0.909; Creatinine).

**Figure 8.**
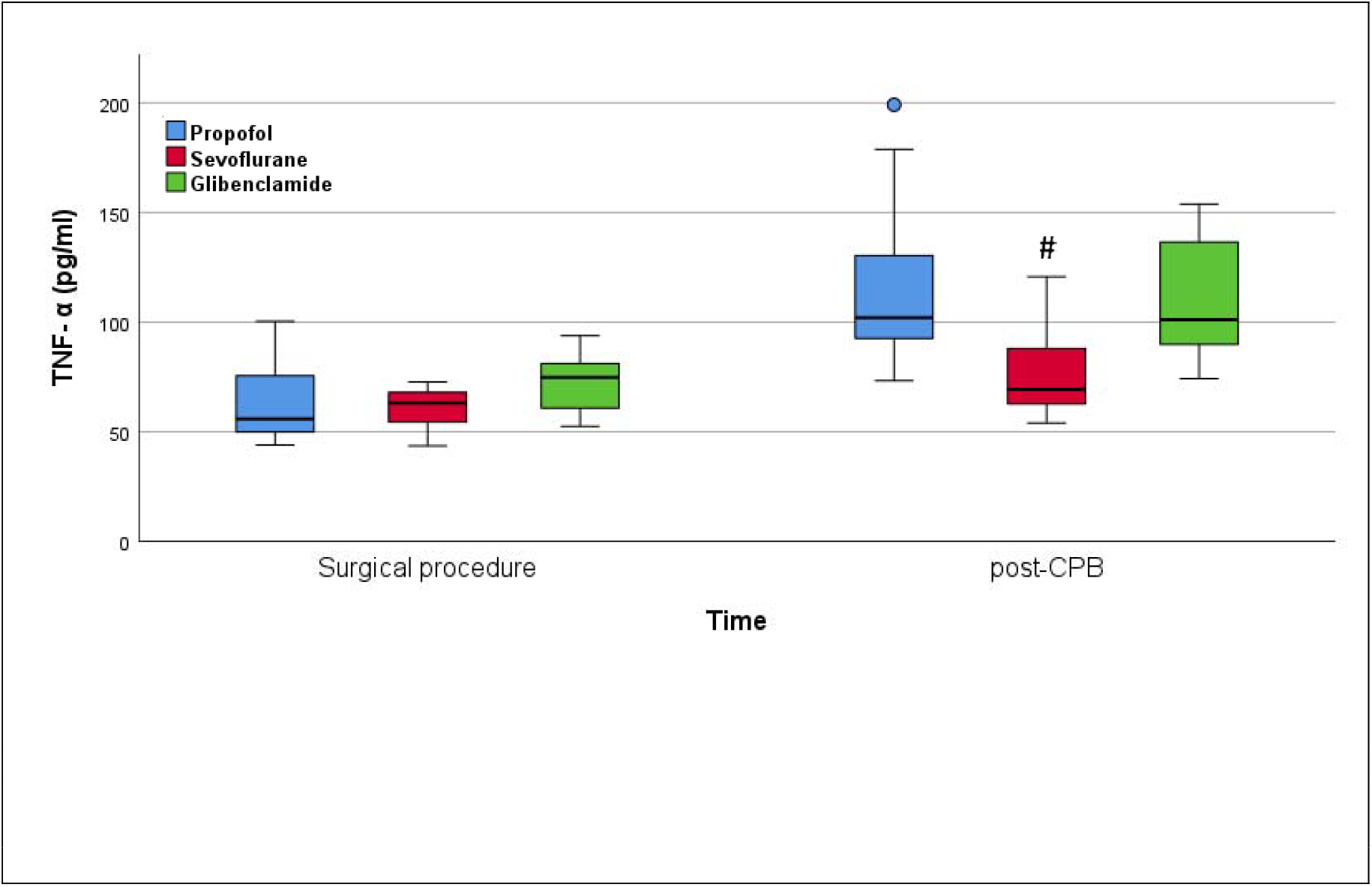
TNF-α: Concentration of TNF-α at the beginning and end of experiment. Surgical procedure resembles baseline values. Post-CPB 90 resembles the end of experiment. Values are expressed as boxplots, continuing the median as a horizontal line inside of the box. The first and third quantile are the edges of the box. Whiskers have a distance about 1.5 interquartile range to the edges. Outliers are plotted as dots. # (p<0.05) represents significant differences between sevoflurane and control group at the respective time steps (*Kruskal-Wallis test with Dunn-Bonferroni correction*).

## Discussion

Our results suggest that sevoflurane may prevent the occurrence of the early onset VS after CPB. No VS effects were seen in the SG with stabile norepinephrine doses at all-time. In this study significant lower levels of TNF-α in the SG compared to control at the end of experiment are shown. This anti-inflammatory effect is well known ^23–25^. In the pathophysiology of the VS cytokine storm leading to NO release is discussed as the main cause of hypotension ^13, 14^. The lower levels of TNF-α in this group could be an explanation for the rather low doses of norepinephrine required to maintain a stable MAP.

Further benefits in hemodynamics were seen as a trend towards higher CI and a physiological HR, this was similar to other studies ^26^. The higher GEF was in one of four measurements significant. With more frequent PICCO^®^ measurements, a significance of the values could underpin the assumption of better cardiac output. In comparison sevoflurane application in a porcine model of Haraldsen et al. showed significantly higher CI values ^9^. In the setting of cardiovascular surgery, sevoflurane is known to have a cardioprotective effect regarding the tolerance of ischemia. This could be another benefit in high risk patients ^27^.

A supposed nephroprotective effect can be shown by the significantly increased urine output in the SG. However, this effect is not reflected in the laboratory parameters.

Even though sevoflurane was shown to be a safe anesthetic with clear advantages in terms of hemodynamics and avoidance of VS, the current climate change must be taken into account when choosing the therapy. Volatile anesthetics may account for 30% of the total hospital emissions ^28^ as greenhouse gases. Therefore, the choice of the right anesthesia should depend on the risk of a patient to develop a VS after complex cardiac surgery.

The application of glibenclamide had multiple, severe effects on the hemodynamic parameters. Through the increase of MAP in a short time, it was possible to reduce the running norepinephrine dose to a minimum. Multiple studies used 10 mg/kg as a bolus injection dose with the same continuous dose over time ^18, 20^. This experience was used to visualize an effect of the study drug that was as clear as possible. In the setting of cardiovascular surgery, it is necessary to find the optimal doses to prevent hypertensive moments shortly after surgery. Early VS was prevented by the application of the drug. 15 min after application, there was no significant difference in MAP between both groups. This phenomenon was seen in other studies as well ^18, 29^. Nevertheless, norepinephrine doses stayed significantly lower until the end of experiment compared to the CG, which could be the effect of continuous application. After initial bolus administration, another effect of the continuous infusion were the significantly higher values of SVRI, not only in the first 15 min but also until the end of experiment. The clear increase in blood pressure can be adequately explained by an increase in vascular tension and thus afterload, which is represented by the SVRI. In contrast, in the CG, higher levels of norepinephrine were not able to generate a similar SVRI over the same period of time. This suggests that glibenclamide has an α-1 receptor independent effect on the vascular tone. In addition, synergistic effects of glibenclamide and norepinephrine are known through a study in a porcine hemorrhagic shock model, where glibenclamide resulted in an improved response of norepinephrine to the vascular tone ^30^.

A further observation is the lowering of the heart rate after administration of glibenclamide as well as a reduction in the CI demonstrated by the PiCCO^®^ measurements. Our observations suggest that this effect is not caused by myocardial damage but rather the return to normal circulatory conditions from initially hyperdynamic conditions after intramuscular premedication with ketamine. It seems as if there is a parasympathetic autoregulation of the heart (negatively inotropic and chronotropic), as the need for cardiac output has decreased. The fact that HR came back to pre-CPB values 25 min after and CI stayed significantly lower for the rest of the experiment suggests that these factors are independent and lower CI is caused by less stroke volume of the left ventricle.

Interestingly, lactate showed significantly higher levels in the GG at the end of experiment.

Due to the fact that there are no significant differences in creatinine values and CKMB between groups, kidney or myocardial ischemia seems unlikely, but a laboratory chemical change may not have developed until the measurement. We further do not assume that myocardial damage occurred because functional parameters such as dPmx did not show significant differences between groups. An autopsy of the animals at the end of the experiment did not show any signs of macroscopic organ ischemia in any group. One explanation for the increased lactate levels could be an interruption of the mitochondrial respiration. This phenomenon is described in studies in which glibenclamide lead to a loss of cellular ATP and blocked the mitochondrial K_ATP_. This effect was concentration-dependent and were only observed in high dose administrations ^31, 32^. Another explanation is that due to higher SVRI and constriction of arteries and arterioles there might be a reduced perfusion in the capillary bed. Further studies are needed to prove the dose of glibenclamide that leads to a sufficient rise of MAP and SVRI that has no side effects in the sense of high lactate levels.

## Conclusion

In the porcine model glibenclamide was able to break through an early VS after CPB significantly raising MAP and SVRI and reduced the need of norepinephrine. Sevoflurane narcosis significantly reduced the occurrence of VS and might therefore be beneficial in high-risk patients.

## Acknowledgements

All Authors have read and are aware of all potential publication charges as listed in the “Cost to Authors” section of the Author Instructions.

## Non-standard abbreviations and acronyms

CPB: Cardiopulmonary Bypass
VS: Vasoplegic syndrome
ATP: Adenine triphosphate
TIVA: Total intravenous anesthesia
VA: Volatile anesthetics
ACC: Aortic Cross Clamping
SG: Sevoflurane Group
CG: Propofol (control) Group
GG: Glibenclamide Group
post-CPB: Pre cardiopulmonary bypass
PCA: Pulse contour analysis
TD: Transpulmonary thermodilution
HLM: Heart-lung machine

## Supplemental Material

Figure 1

Figure 2

